# Estimation of the Time-Varying Reproduction Number of COVID-19 Outbreak in China

**DOI:** 10.1101/2020.02.08.20021253

**Authors:** Chong You, Yuhao Deng, Wenjie Hu, Jiarui Sun, Qiushi Lin, Feng Zhou, Cheng Heng Pang, Yuan Zhang, Zhengchao Chen, Xiao-Hua Zhou

## Abstract

**Background:** The 2019-nCoV outbreak in Wuhan, China has attracted world-wide attention. As of February 11, 2020, a total of 44730 cases of novel coronavirus-infected pneumonia associated with COVID-19 were confirmed by the National Health Commission of China.

**Methods:** Three approaches, namely Poisson likelihood-based method (ML), exponential growth rate-based method (EGR) and stochastic Susceptible-Infected-Removed dynamic model-based method (SIR), were implemented to estimate the basic and controlled reproduction numbers.

**Results:** A total of 71 chains of transmission together with dates of symptoms onset and 67 dates of infections were identified among 5405 confirmed cases outside Hubei as reported by February 2, 2020. Based on this information, we find the serial interval having an average of 4.41 days with a standard deviation of 3.17 days and the infectious period having an average of 10.91 days with a standard deviation of 3.95 days.

**Conclusions:** The controlled reproduction number is declining. It is lower than one in most regions of China, but is still larger than one in Hubei Province. Sustained efforts are needed to further reduce the *R*_*c*_ to below one in order to end the current epidemic.

## 1. Introduction

On December 29, 2019, Wuhan, the capital city of Hubei Province in Central China, has reported four cases of pneumonia with unknown etiology. Since then, the outbreak has rapidly worsened over a short span of time and has received considerable global attention. On January 7, 2020, the pathogen of the current outbreak was identified as a novel coronavirus (2019-nCoV), and its gene sequence was quickly submitted to WHO (The coronavirus was renamed COVID-19 by WHO on February 12).^1,2^ On January 30, WHO announced the listing of this novel coronavirus-infected pneumonia (NCP) as a “public health emergency of international concern”. As of February 11, 2020, the National Health Commission (NHC) of China had confirmed a total of 44730 cases of novel coronavirus-infected pneumonia (NCP) associated with the COVID-19 in Mainland China, including 1114 fatalities and 4771 recoveries.

Since January 19, 2020, strict containment measures, including travel restrictions, contact tracing, entry or exit screening, non-hospital isolation, quarantine, awareness campaigns and others, have been implemented by the Wuhan Government and quickly adopted by other cities within China with the aim to minimize virus transmission via human-to-human contact. Similar measures were previously employed in China in 2009 to tackle the outbreak of H1N1 including mandatory quarantine of anyone who has had close contact with confirmed patients.

This article investigates the change in the basic reproduction number *R*_0_ and controlled reproduction number *R*_*c*_ since the outbreak of COVID-19. We have found that the estimated controlled reproduction numbers *R*_*c*_ in all different regions are significantly smaller compared with the basic reproduction numbers *R*_0_, but *R*_*c*_ is still greater than one in Hubei Province.

## 2. Data

Data were collected from provincial/municipal health commissions in China as well as through ministry of health in other countries and regions with details of each confirmed case including case ID, region, age, gender, date of symptom onset, date of diagnosis, history of traveling to or residing in Wuhan, and, if any, related remarks such as contact identification, cases and case-related information. In addition, the collected dataset also contains date of infection and chains of transmission of infection which can be inferred from travel or residency history in Wuhan and other relevant information, if available, as follows:

1. If the individual has not been to Hubei Province recently, but were exposed within a four-day period (i.e., the individual has had contact with a confirmed case of NCP on a certain day), then the corresponding date of infection is inferred as the middle of the exposure period;
2. If the individual has travelled to Hubei Province but has returned within four days, then the date of infection is inferred as the middle of the travelling period;
3. If the individual has not been to Hubei Province recently, but in close contact with an imported case from Hubei, then the individual is identified to be infected by this imported case;
4. If the individual has not been to Hubei Province recently, but in close contact with a local case who was clearly infected before that individual, then this individual is identified to be infected by the corresponding local case.

Note: if the individual has been to Hubei Province, the transmission history would not be recorded despite the existence of contact tracing information.

## 3. Inference about the Serial Interval and Infectious Period

In this study, serial interval is defined as the time difference between dates of infection of successive cases in a chain of transmission (different textbooks may have different definitions). Infectious period is the duration of which an infected individual can transmit pathogens to a susceptible host. In this study, infectious period is defined as the time difference between date of infection and date of diagnosis as there is strong evidence showing that a diseased individual remains contagious even during the incubation period, and would be immediately isolated upon positive diagnosis hence losing the transmissibility. Both are key quantities that depict an epidemic and are essential to estimate the basic/controlled reproductive number, *R*_0_ / *R*_*c*_. Among 139 chains of transmission identified from 5405 confirmed cases outside Hubei as recorded by February 2, 2020, none of them have their dates of infection acquired, but 71 of them have their dates of symptoms onset available.

Hence, the corresponding serial intervals were approximated by the differences in dates of symptom onset rather than the actual dates of infection, see Figure 1.

**Figure 1:**
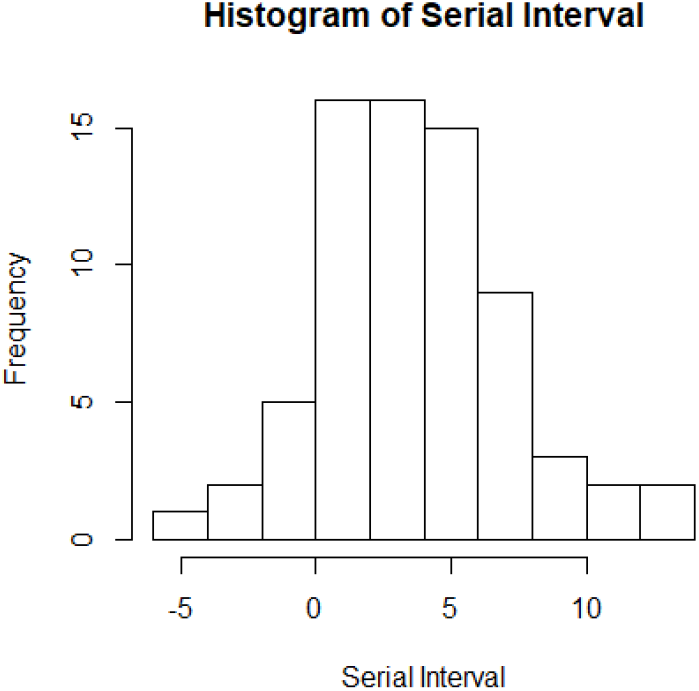
Histogram of serial interval with the average of 4.27 days before correction.

We can see that some serial intervals are negative which is certainly impossible by definition. However, noting that the serial intervals were approximated from the dates of symptoms onset, this suggests that the negative values could be caused by different lengths of incubation period between individuals. Here a simple correction is implemented by resetting the negative values to zeros. This shifts the average of the serial intervals to 4.41 days and the standard deviation to 3.17 days after corrections, see table 1. Note that the serial interval of SARS-nCoV in Hongkong was 8.4 days on average.^3^ In addition, a total of 67 cases in the collected data were able to identify the corresponding dates of infection. Figure 2 plots the histogram of infectious period while Table 2 shows the numerical summary.

**Table 1:**
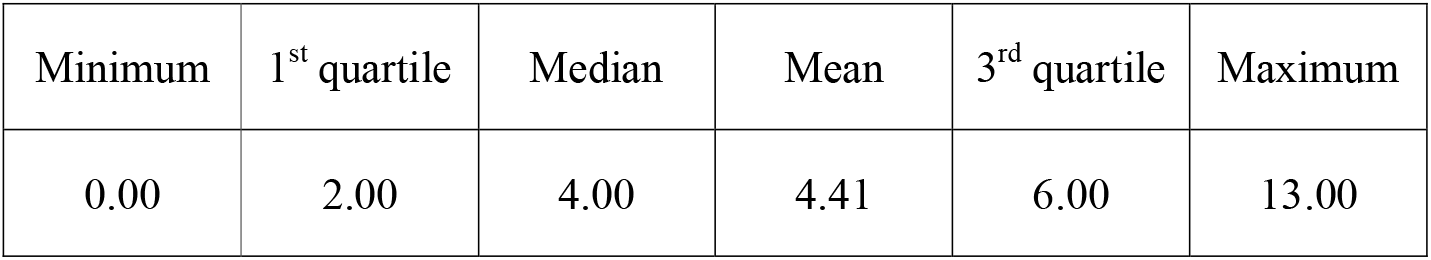
numerical summary of serial intervals.

**Table 2:** numerical summary of infectious period.

| Minimum | 1<sup>st</sup> quartile | Median | Mean | 3<sup>rd</sup> quartile | Maximum |
| --- | --- | --- | --- | --- | --- |
| 2.00 | 8.00 | 11.00 | 10.91 | 13.00 | 25.00 |

**Figure 2:**
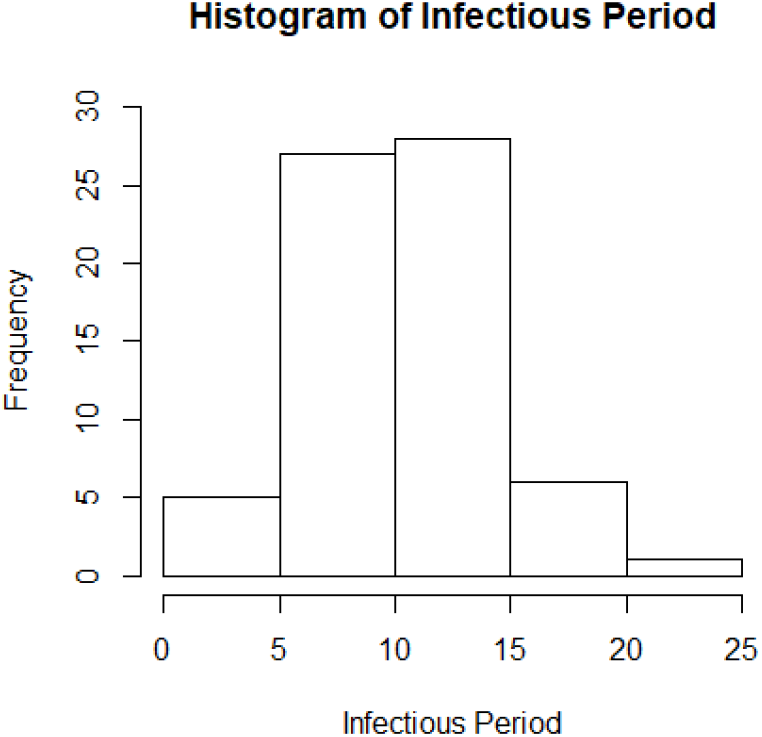
Histogram of infectious period with the average of 10.91 days.

We found that there were no significant demographical differences between the subset of cases used to estimate serial interval and infectious period and the cases in the full dataset. Therefore, the inference made on serial interval and infectious period based on the corresponding subsets should be able to represent the full dataset.

## 4. Estimation of Basic/Controlled Reproduction Number

### Definition

The reproduction number *R*_0_is defined as the (average) number of new infections generated by one infected individual during the entire infectious period in a fully susceptible population.^4^ It can be also understood as the average number of infections caused by a typical individual during the early stage of an outbreak when nearly all individuals in the population are susceptible. The basic reproduction number reflects the ability of an infection spreading under no control. When the size of susceptible population is limited, the quantity, effective reproduction number *R*_*e*_, is used instead of *R*_0_. Similarly, the quantity, controlled reproduction number *R*_*c*_, should be used to describe the ability of disease spreading when interventions (such as quarantine, isolation, or traffic control) are taking place. Hence a good measure of any intervention is to reduce *R*_*c*_. Note that the disease will decline and eventually die out if *R*_*c*_ *≤1*.

### Methods

The basic reproduction number can be estimated through a variety of models.^5^ In this section, we have compared three most popular estimates of *R*_0_/*R*_*c*_ as shown below.

#### (1) Poisson Likelihood-based (ML) method

Let *N*_*t*_ be the number of reported new confirmed cases on day *t*. Suppose that the serial interval has a maximum of *k* days and the number of new cases generated by an infected individual is assumed to follow a Poisson distribution with parameter *R*_0_.^6^ The probability that the serial interval of an individual in *1* days is *w*_*j*_, which can be estimated from the empirical distribution of serial interval or by setting up a discretized Gamma prior on it. Thus, the likelihood function can be reduced into a thinned Poisson

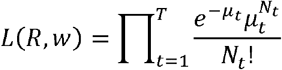

where

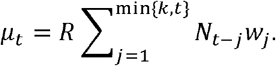

The reproduction number R can be estimated by maximizing the likelihood function. Note that if the empirical distribution of serial interval is used or *w*_*j*_, are given, then

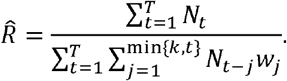

#### (2) Exponential growth rate-based (EGR) method

At the early period of an epidemic, the number of infected cases rises exponentially. Suppose the exponential epidemic growth rate (Malthusian coefficient) is R, which can be estimated by fitting a least square line to the daily number of reported new confirmed cases in a log-scale, namely, *log* (*N*_*t*_). Let *f*_*G*_(*t*) denote the probability density function of serial interval. Hence the reproduction number can be calculated according to the Euler-Lotka equation in a moment generating form^7^

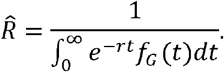

#### (3) Stochastic dynamic model-based method

Here we consider a stochastic Susceptible-Infected-Removed (SIR) model rather than a standard deterministic one. The major advantage of using a stochastic dynamic model is that it affords improved accounting for real variabilities and increases opportunity for quantifying uncertainties.^8^ Let *S(t), I(t)* and *R(t)* denote the number of susceptible, infectious and recovered population at time *t* respectively, and note that *N= s(t) + 1(t) + R(t)*. Suppose that the infectious period of an individual is a random variable *T∼ Exp(γ)*, then the reproduction number *R= β(T) = β/ γ*, where *γ* and *β* are the recovery rate and transmission rate respectively in the system of ordinary differential equation (ODE) below,

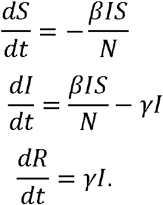

The maximum likelihood method is used to estimate model parameters where the likelihood is obtained by sequential Monte Carlo method, and parameters are estimated using the Iterated Filtering algorithm (IF2)^9^ implemented as `mif` in the `R` package `pomp`^10^ with *s(0)* equals the population of the region, *R(0) = 0, I(0)* is 10 times the average number of confirmed cases from Day 0 to Day 7 and *γ = 10*.*91* obtained from the collected data described ahead.

### Results

In this section we have estimated the basic reproduction number *R*_0_ and the controlled reproduction number *R*_*c*_. Since January 19, 2020, various containment measures have been strictly implemented, especially after the State Council agreed to include NCP into the Management of the Infectious Diseases Law and the Health and Quarantine Law on January 20. Based on an average10.91-day infectious period estimate from our collected data, we expect a flatter rate of increment starting on January 29. Figure 3 plots the number of daily new cases in a log-scale against date, and, as anticipated, the trend supports our guess.Therefore, the quantities *R*_0_ and *R*_*c*_ are estimated based on collected data in two separate periods, i.e., from January 21 (the starting date of daily updates of confirmed cases nationwide) to January 28, and from January 29 to February 5 (the end date of this study) respectively.

**Figure 3:**
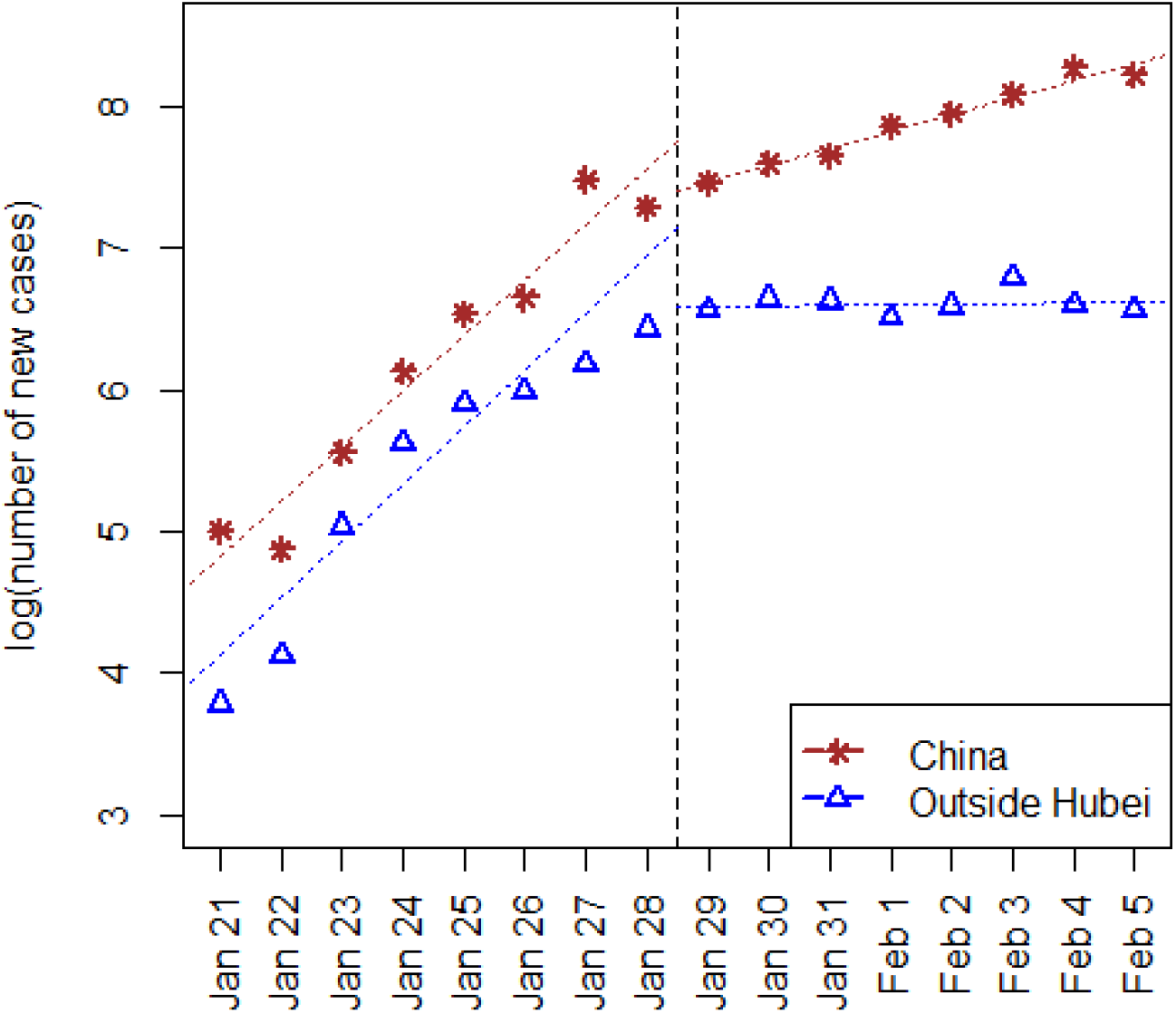
Visualization of daily numbers of new confirmed case along with date, as obvious change of rate occurred on January 29 as expected.

The estimates of *R*_0_ and *R*_*c*_ by Poisson likelihood (ML) and exponential growth rate (EGR) in selected regions of China are listed in Table 3 and Table 4. Despite the disagreement between different estimation methods, all three methods indicate notable reductions from *R*_0_ to *R*_*c*_ which suggests an improvement in the current situation. This is possibly due to the effective interventions and prompt actions by the local and central governments to minimize further spreading. We also notice that EGR yields smaller estimates of *R*_*c*_ compared to other methods. This might be because the number of infected patients does not grow exponentially after such strict containment measures.

**Table 3:** Estimates and 95% confidence intervals of basic reproduction number in some selected provinces (or cities) of China, from Jan 21 to Jan 28, 2020.

|  | ML | EGR | SIR |
| --- | --- | --- | --- |
| China | 3.024 (2.020, 4.422) | 3.738 (2.502, 6.006) | 5.4 (4.5, 6.2) |
| Hubei | 3.334 (2.118, 5.056) | 3.758 (2.052, 7.440) | 5.5 (4.2, 6.8) |
| Others | 2.681 (1.754, 3.941) | 3.687 (2.281, 6.474) | 5.1 (3.9, 6.3) |
| Beijing | 2.278 (1.436, 3.281) | 2.164 (1.129, 4.116) | 2.3 (1.1, 3.8) |
| Shanghai | 2.144 (1.373, 3.002) | 1.610 (0.636, 3.441) | 2.4 (1.0, 3.8) |
| Guangdong | 2.488 (1.692, 3.493) | 2.691 (1.337, 5.335) | 3.7 (2.7, 4.9) |
| Zhejiang | 2.506 (1.649, 3.626) | 3.132 (1.335, 6.958) | 5.0 (3.3, 7.0) |
| Hunan | 2.767 (1.795, 4.086) | 4.872 (2.171, 11.136) | 5.3 (4.3, 7.0) |
| Henan | 2.888 (1.783, 4.400) | 5.452 (2.650, 11.771) | 6.4 (3.5, 10.2) |

**Table 4:** Estimates and 95% confidence intervals of controlled reproduction number in some selected provinces (or cities) of China, from Jan 29 to Feb 5, 2020.

|  | ML | EGR | SIR |
| --- | --- | --- | --- |
| China | 2.282 (1.726, 2.985) | 1.652 (1.457, 1.906) | 2.3 (2.1, 2.5) |
| Hubei | 2.503 (1.908, 3.321) | 1.935 (1.642, 2.367) | 2.8 (2.5, 3.1) |
| Others | 1.840 (1.465, 2.332) | 1.071 (0.915, 1.263) | 1.2 (1.1, 1.4) |
| Beijing | 2.260 (1.644, 3.013) | 1.619 (0.861, 2.986) | 2.1 (1.0, 3.3) |
| Shanghai | 1.876 (1.407, 2.446) | 1.073 (0.673, 1.616) | 1.2 (0.7, 2.0) |
| Guangdong | 1.899 (1.480, 2.482) | 1.131 (0.718, 1.712) | 1.2 (0.8, 1.8) |
| Zhejiang | 1.480 (1.199, 1.838) | 0.654 (0.343, 1.014) | 1.0 (0.4, 1.7) |
| Hunan | 1.698 (1.333, 2.151) | 0.963 (0.677, 1.302) | 1.3 (1.0, 1.8) |
| Henan | 2.091 (1.597, 2.657) | 1.403 (1.054, 1.941) | 1.5 (1.1, 2.0) |

Furthermore, the time-varying controlled reproduction number *R*_*c*_(*t*) can be estimated through the Poisson likelihood (ML) method where *t* is from Feb 1 to Feb 10, 2020. For each Day *t*, the number of daily reported new cases from Day *t-7* to Day *t* is used to estimate 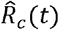. Figure 4 plots the estimated controlled reproduction number 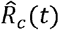 along with its 95% CI for selected regions of China. Note that the estimated 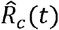 reflects the average spreading ability of the epidemic in a short period prior to Day *t*. As a result, the real-time *R*_*c*_(*t*) might be overestimated if the general trend of *R*_*c*_(*t*) is declining.

**Figure 4:**
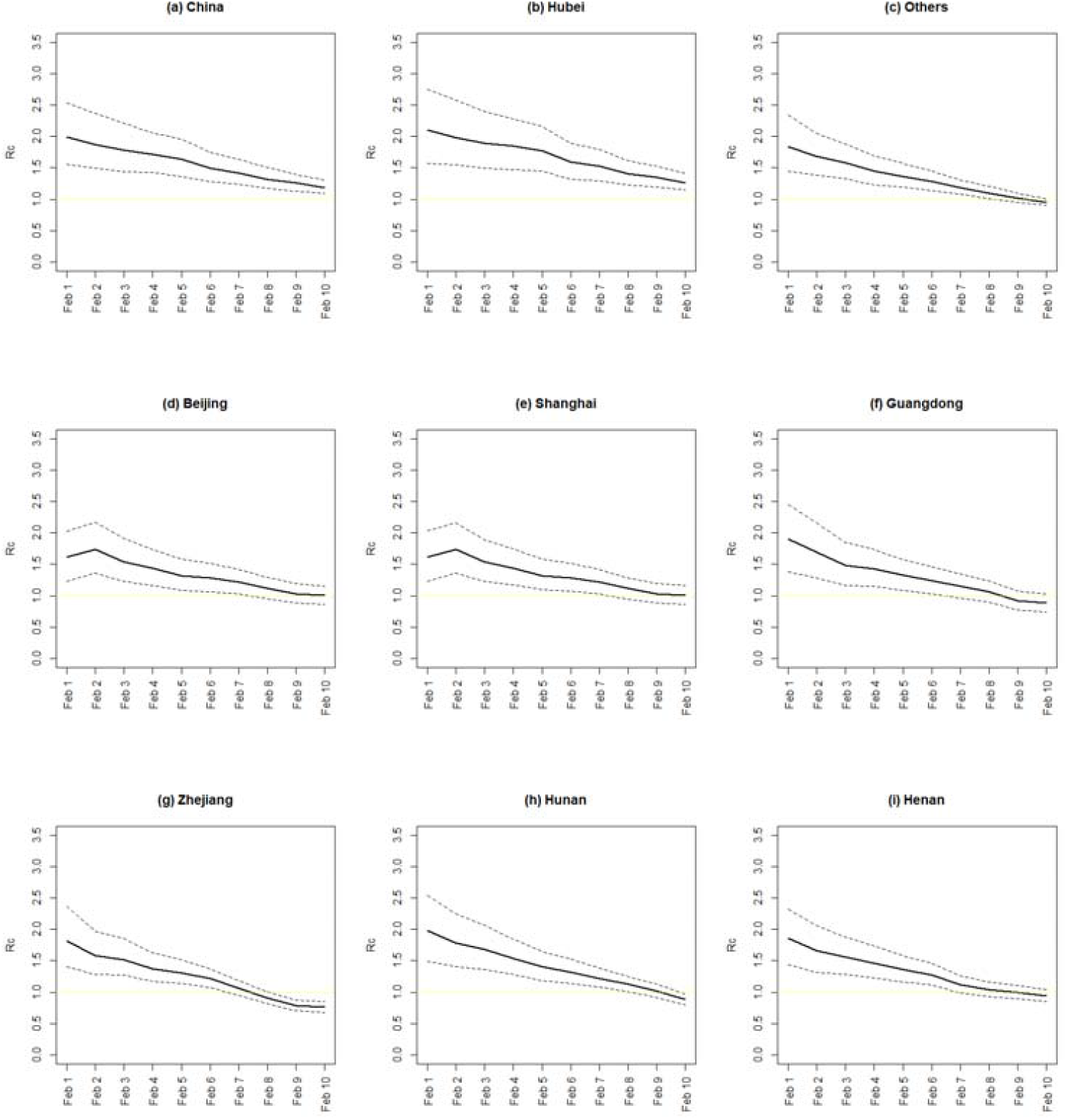
The estimated controlled reproduction number in (a) China, (b) Hubei, (c) Other provinces except Hubei, (d) Beijing, (e) Shanghai, (f) Guangdong, (g) Zhejiang, (h) Hunan, and (i) Henan. The dashed line is the 95% confidence interval.

## 5. Conclusion

Despite the continuous increase in new confirmed cases on a daily basis, the estimated controlled reproduction numbers *R*_*c*_ produced by all three methods in all different regions are significantly smaller compared with the basic reproduction numbers *R*_0_. As discussed in Section 4, the real-time controlled reproduction number may be even lower than the estimated values in Figure 3. Nonetheless, additional effort is needed to further reduce *R*_*c*_ below one in Hubei Province.

## 6. Discussion

The dataset used in this study is based on the confirmed cases reported by NHC China. However, during the period of data collection, the official guidelines for diagnosis and treatment were updated four times. The criteria of confirmation have evolved from the original “whole genome sequencing of the respiratory excretion” to “positive viral nucleic acid results by the RT-PCR of the respiratory excretion or viral gene sequence”, and, most currently, the inclusion of positive nucleic acid results of the blood sample. In the meanwhile, the confirmation process is simplified by removing the accreditation process by the national expert committee for confirmed cases. The fourth edition granted the accrediting authority to the municipalities.^11^ In addition, the medical resources in Hubei Province especially in Wuhan have been enhanced remarkably. All of these changes might result in a temporary surge of confirmed cases and lead to an overestimation of *R*_*c*_, especially in Hubei Province.

Furthermore, the current containment measures mainly aim to cut the transmission from human to human via droplets of respiratory. However, other transmission pathways, including fecal-oral transmission and aerosol transmission, could not yet be excluded based on current evidence. If other transmission mechanisms do exist, the *R*_*c*_ values would remain high in the future unless further measures would intersect these transmission pathways.

## Data Availability

All data are collected from the website of China CDC.

http://2019ncov.chinacdc.cn/2019-nCoV/

## Acknowledgments

We thank Taojun Hu, Xueqing Liu and Yuying Li from School of Public Health, Peking University for assistance of data collection.

## Notes

### Competing Interest Statement

The authors have declared no competing interest.

### Funding Statement

No funding.

